# Estimate the incubation period of coronavirus 2019 (COVID-19)

**DOI:** 10.1101/2020.02.24.20027474

**Authors:** Ke Men, Xia Wang, Li Yihao, Guangwei Zhang, Jingjing Hu, Yanyan Gao, Henry Han

## Abstract

**Motivation:** Wuhan pneumonia is an acute infectious disease caused by the 2019 novel coronavirus (COVID-19). It is being treated as a Class A infectious disease though it was classified as Class B according to the Infectious Disease Prevention Act of China. Accurate estimation of the incubation period of the coronavirus is essential to the prevention and control. However, it remains unclear about its exact incubation period though it is believed that symptoms of COVID-19 can appear in as few as 2 days or as long as 14 or even more after exposure. The accurate incubation period calculation requires original chain-of-infection data that may not be fully available in the Wuhan regions. In this study, we aim to accurately calculate the incubation period of COVID-19 by taking advantage of the chain-of-infection data, which is well-documented and epidemiologically informative, outside the Wuhan regions.

**Methods:** We acquired and collected officially reported COVID-19 data from 10 regions in China except for Hubei province. To achieve the accurate calculation of the incubation period, we only involved the officially confirmed cases with a clear history of exposure and time of onset. We excluded those without relevant epidemiological descriptions, working or living in Wuhan for a long time, or hard to determine the possible exposure time. We proposed a Monte Caro simulation approach to estimate the incubation of COVID-19 as well as employed nonparametric ways. We also employed manifold learning and related statistical analysis to decipher the incubation relationships between different age/gender groups.

**Result:** The incubation period of COVID-19 did not follow general incubation distributions such as lognormal, Weibull, and Gamma distributions. We estimated that the mean and median of its incubation were 5.84 and 5.0 days via bootstrap and proposed Monte Carlo simulations. We found that the incubation periods of the groups with age>=40 years and age<40 years demonstrated a statistically significant difference. The former group had a longer incubation period and a larger variance than the latter. It further suggested that different quarantine time should be applied to the groups for their different incubation periods. Our machine learning analysis also showed that the two groups were linearly separable. incubation of COVID-19 along with previous statistical analysis. Our results further indicated that the incubation difference between males and females did not demonstrate a statistical significance.

## Introduction

Novel coronaviruses (COVID-19), which was found in Wuhan, China in December 2019 presents an acute public health threat to the whole world [1-2]. The new virus is different from known coronaviruses such as SARS and MERS, though they share some similar respiratory illness symptoms such as fever, cough, or/and shortness of breath [2]. It is believed to root from the animal but spreads from person-to-person. The COVID-19 spread even shows that persons without any symptoms or clinically negative in infection can still spread it to others. There is no official vaccine or antiviral drug available up to now (Jan 30, 2020) to treat COVID-19 infected patients [3-4]. The outbreak of COVID-19 infection is forcing China and many countries to take harsh protection policies. More than eight-thousands of infections have been reported in China and more than a dozen of countries until Jan 30, 2020. More than 15 cities including Wuhan have been quarantined to halt the spread of the COVID-19. It is expected that millions of people can be on lockdown because of COVID-19. WHO declared the coronavirus outbreak a global health emergency on Jan 30, 2020.

It is essential to know the accurate incubation period of COVID-19 for the sake of deciphering dynamics of its spread. The incubation period is the time from infection to the onset of the disease. It provides the foundation for epidemiological prevention, clinical actions, and drug discovery. Different viruses have different incubation periods that determine their different dynamics epidemiologically. The incubation period of H7N9 (Human Avian Influenza A) is about 6.5 days, but the incubation period for SARS-CoV is typically 2 to 7 days [5-6]. However, it remains unclear about its exact incubation period of COVID-19, although WHO estimates it is between 2 to 14 days after exposure [8]. It can be difficult to estimate the incubation period of COVID-19 by using original chain-of-infection data that may not be fully available in the Wuhan regions. Or data may lack meaningful exposure history. Furthermore, it is also unknown whether the incubation time will show some statistically significant with respect to Age and Gender. In this study, we aim to accurately estimate the incubation period of COVID-19 by taking advantage of datasets with a well-documented history of exposure. Our results show the incubation mean and median of COVID-19 are 5.84 and 5.0 days respectively and there is a statistical significance with the role of gender. However, the incubation periods of the groups with age>=40 years and age<40 years show a statistically significant difference. Our machine learning analysis also shows that the two groups are linearly separable that demonstrate a clear boundary in knowledge discovery visualization.

## Methods

### Data collection and preprocessing

We collected a dataset with 59 officially confirmed COVID-19 cases from 10 regions in China except for Hubei province, the assumed origin of the virus. The patient data was dated from Dec 29, 2019, to Feb 5, 2020. We only involved the officially confirmed cases with a clear history of exposure and time of onset in data collection. We exclude those without relevant epidemiological descriptions, working or living in Wuhan for a long time, or hard to determine the possible exposure time.

Data collected for this study included region, age, gender, exposure history, and illness onset. For those cases whose incubation periods locate in an interval [*x*_1_, *x*_2_], we use its midpoint 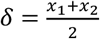 to represent its incubation period. For example, Case no. 2 in our dataset went on a business trip in Wuhan on Jan 12^th^, 2020 and returned to Shaanxi on Jan 15^th^, 2020, but had fever symptoms on Jan 20^th^ 2020. The incubation will be calculated as 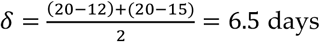. More details about the dataset can be found in the Result section.

### Monte Carlo simulation for incubation median and mean estimation

We propose a Monte Carlo simulation approach that takes advantage of bootstrap techniques to estimate incubation median and mean estimation for the small sample with 59 cases. It is more data-driven compared to traditional parametric approaches to handle parameter estimation for small datasets. The proposed Monte Carlo simulation assumes we have a collected small incubation dataset *X*. We generate a large incubation sample *S*_*i*_ by concatenating *n*=1000 randomly sampled incubation segments *u*_*ik*_ each of which contains at least *l* entries in the interval [*l*_1_, *l*_2_] drawn independently from the existing dataset *X*, i.e., 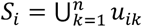 Then the large incubation sample median is calculated: *m*_*i*_ = *median*(*S*_*i*_) + *ρ* × *ϵ*(0,1),where *ϵ*(0,1) is added Gaussian noise and *ρ* ∈ [0,1] is a variance control parameter in simulation. Such aprocedure is repeated *N* times, the population median 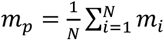. The estimated standard deviation *σ*_*p*_ is calculated as the standard deviation of the median sequence *m*_1_, *m*_2_, … *m*_*n*_. In our simulation, we chose *N* = 100000, *l*_1_ = 1, *l*_2_ = 7, *ρ* = .2 days in simulation and conduct simulations by employing Google Colab with TPU acceleration [9-10]. The confidence level probability is calculated by the ratio *T*/*N*, where *T* counts the times that *m*_*i*_ falls in the interval [*m*_*p*_ − 2*σ*_*p*_, *m*_*p*_ + 2*σ*_*p*_] H in the simulation.

Similarly, we can estimate the population mean by using the same way where the large incubation sample mean is calculated as 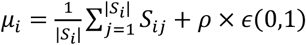 each time. The population mean is estimated as 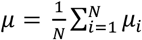 and the estimated standard deviation *ρ*. According to the central limit theorem, the population mean will be subject to the normal distribution. The confidence interval probability is calculated by following the same procedure.

## Result

Our data consists of 34 male cases, 24 female cases and 1 unidentified gender case from ten regions in China. All 59 cases have complete epidemiological descriptions about the history of exposure. The total 57 cases have complete information in age and gender One case from Beijing has unknown age. The mean and standard deviation of age are 41.9 and 13.2 years old. The mode of his dataset is 4.0 with 14 support cases. The minimum and maximum age are 10 and 70 respectively.

Figure 1 summarizes age and incubation time variables with respect to region and gender. The median incubation period of males (5.0 days) is slightly shorter than that of females 5.5 days. The median ages of male and female groups are 41 and 42 years old although their age distributions are quite different. The minimum and maximum incubation periods are 1 and 15 days respectively. Among all 10 regions, Shaanxi and Guangdong have 18 and 14 cases that are the largest and 2^nd^ largest subgroups whose median incubation periods are 5.0 and 5.5 respectively.

**Fig 1.**
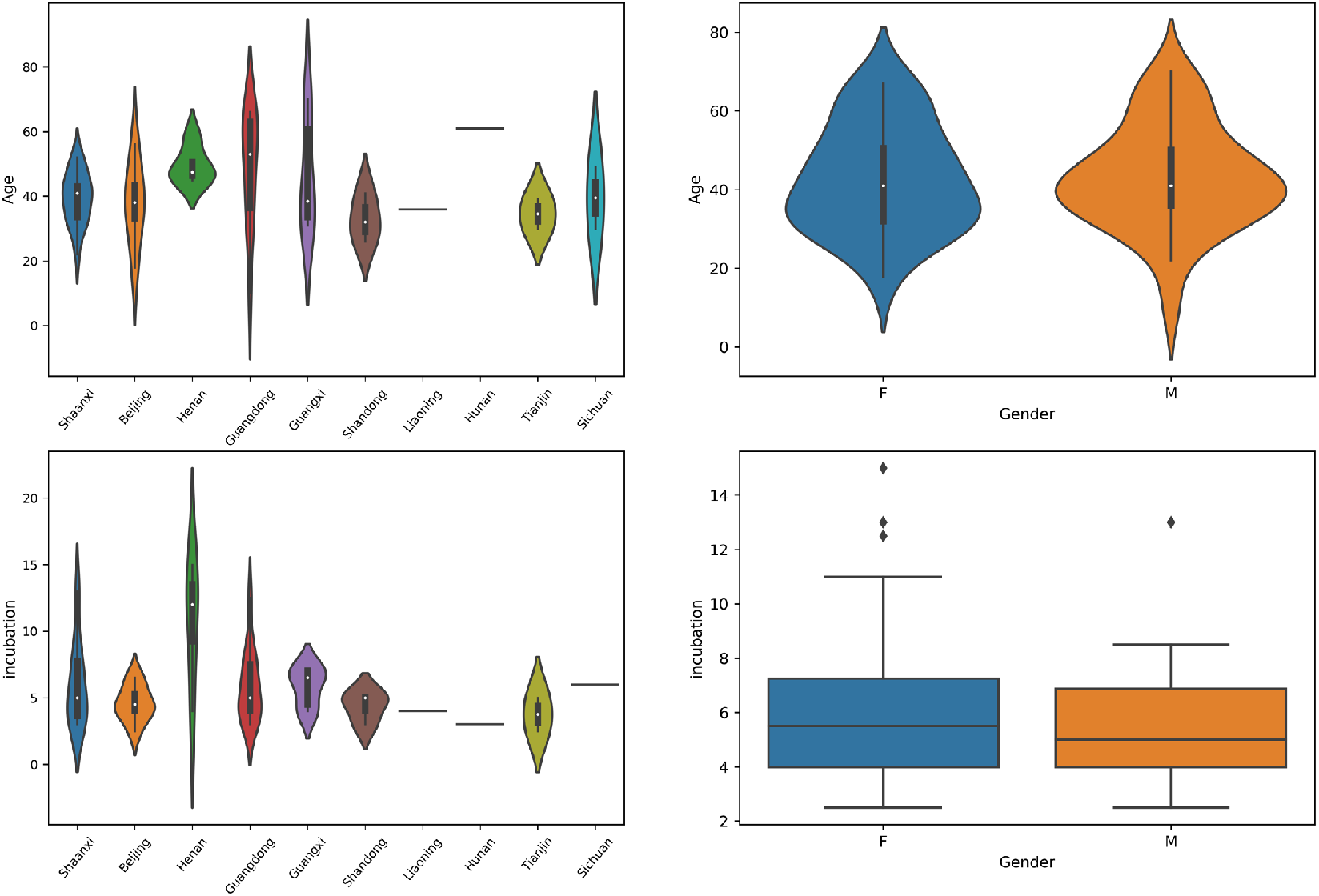
Age and incubation time summary with respect to region and gender.

Table 1 shows incubation statistics for five different groups: Male, Female, Age>=40, Age <40, and All that include all cases. It indicates that the incubation period median and mean of patients more than 40 years old are greater than those of patients less than 40 years old. Similar patterns are also observed for male and female groups. The mean values are always larger than median values for each group suggests the right-skewed distributions of incubation.

**Table 1.** Incubation statistics for different groups

| Group | The number of cases | Incubation statistics |  |  |
| --- | --- | --- | --- | --- |
| | | mean ± std | median | IQR |
| Male | 34 | 5.54±2.57 | 5.0 | 2.88 |
| Female | 24 | 6.33±3.41 | 5.5 | 3.25 |
| Age ≥40 | 32 | 6.73±3.20 | 6.0 | 4.0 |
| Age <40 | 25 | 4.84±2.28 | 4.0 | 1.0 |
| All | 59 | 5.84±2.93 | 5.0 | 3.0 |

Figure 2 illustrates the probability density functions (p.d.f) of ‘male’, ‘female’, and ‘all’ group incubation using Gaussian kernel density estimation on the left plot. The three groups seem to have similar or the same incubation distributions. The right plot in Figure 2 compares the probability function distributions (p.d.f.) of those of the ‘age>= 40 years’ and ‘age < 40 years’ groups. It suggests that the age>=40 group demonstrates very different incubation distributions compared to the age<40 group.

**Fig 2.**
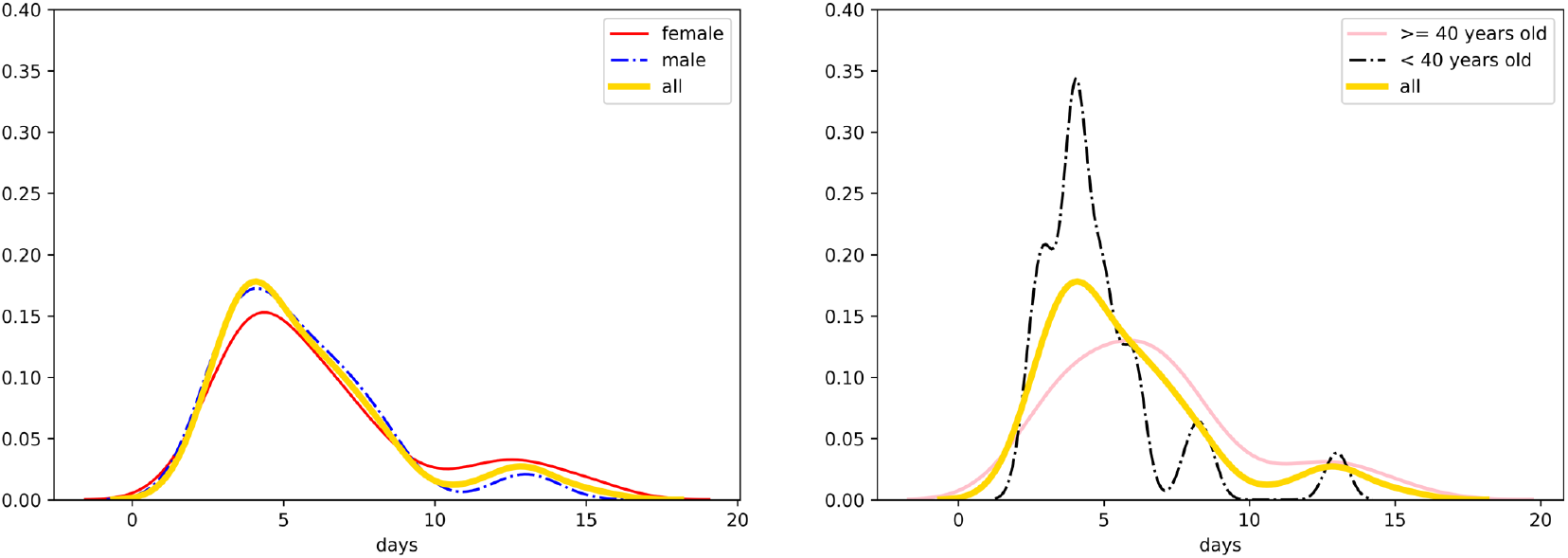
The probability density functions (p.d.f.) under Gaussian kernel density estimation for different groups.

### The distributions of the incubation of COVID-19

The incubation of COVID-19 is not subject to neither of the widely used incubation distributions such as normal, lognormal, Gamma, and Weibull distributions well [12]. We employ Shapiro-Wilk tests rather than Kolmogorov–Smirnov tests to conduct normality tests because we only have 59 records and Shapiro-Wilk tests can do a much better job on small datasets with a sample size of from 3 to 5000 [13]. Table 2 shows the *p*-values under the Shapiro-Wilk tests for normal and lognormal distributions as well as Goodness-of-fit tests for Gamma and Weibull distributions by using the R package ‘*goft*’ [14]. We can reject normal, lognormal, and Weibull distributions strongly under the significance level of 0.05 cutoff [11-12]. Although we can’t reject Gamma distributions for its boundary line *p*-value (0.06086), it can be risky to use it to fit and estimate the distribution of the incubation period under a small sample size. We further employ the maximum likelihood estimates (MLEs) for the parameters of the gamma, conduct the goodness-fit test, and obtain *p*-value=8.6807e-04. As such, we only rely on nonparametric techniques in data analysis rather than use any pre-assumed distributions.

**Table 2.** *p-*values for probability distribution tests

| Distribution | $p$-value |
| --- | --- |
| Normal | 2.009e-06 |
| Lognormal | 0.01396 |
| Gamma | 0.06086 |
| Weibull | 6e-05 |

### The incubation difference between males and females does not demonstrate a statistical significance

Mann-Whitney rank test shows that there are no significant differences between the incubation of males and females with respect to median with *p*-value=0.267 for the null hypothesis *H*_0_: Male and female groups have the same median. Similarly, we employ the Siegal-Tukey test to find the variances of incubation of males and females are at the same level statistically. We have *p*-values: 0.74725, 0.376, and 0.6258 for the corresponding hypotheses respectively: *H*_1_: male and female groups have different incubation variances, *H*_2_: the variance of male incubation > and that of the females, and *H*_3_: the variance of male incubation < that of the females. The results indicate that gender may not be a key factor affecting the incubation period of COVID-19.

### The incubation period of the age<40 group is statistically shorter than that of the age>=40 group

Pearson correlation coefficient analysis shows the R statistics is 0.244 with *p*-value: 0.06758. It suggests that that the incubation period is somewhat correlated with age though not that strong. To verify whether the incubation of the age>=40 group is different from that of the age<40 group statistically, Figure 3 compares their incubation periods groups using different visualization tools. It indicates that the younger group tends to have a shorter incubation period. The variance of their incubation period also seems to be smaller.

**Fig 3.**
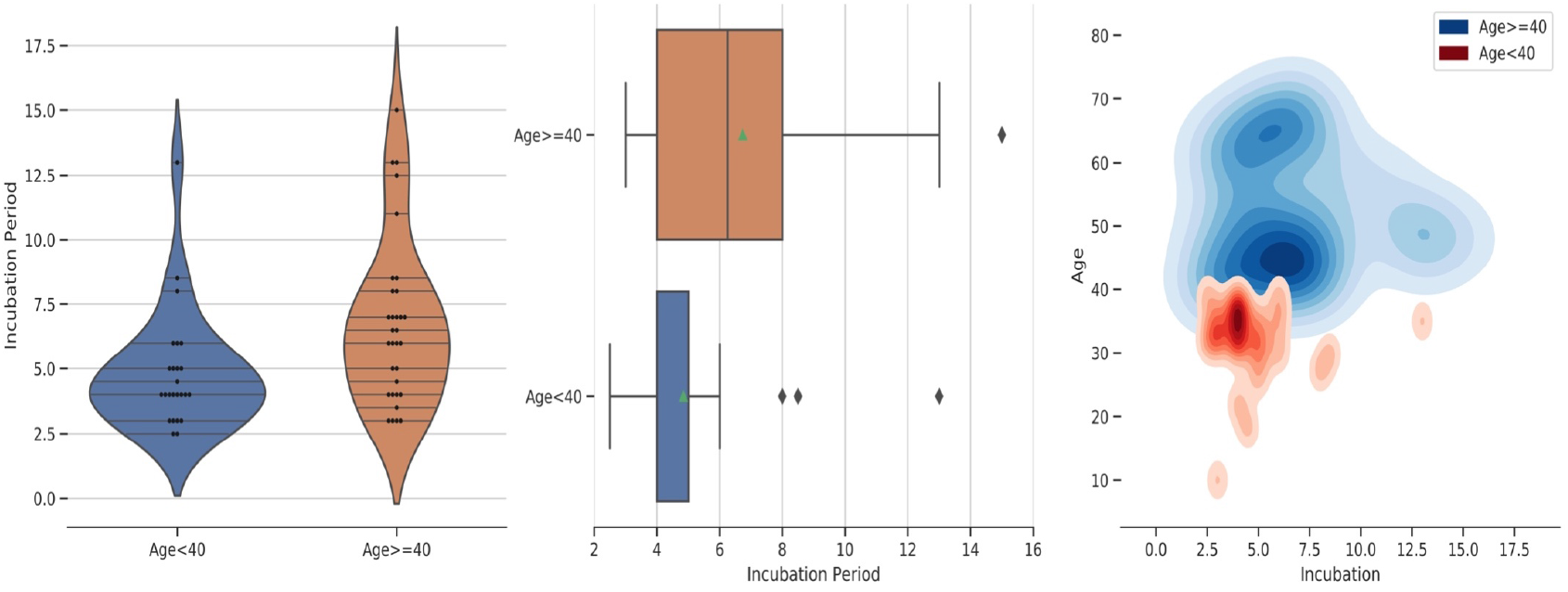
Compare the incubation periods of Age>=40 and Age< 40 groups.

The Mann-Whitney rank test shows that there’s a statistically significant difference between the incubation of age<40 and age>=40 groups with the null hypothesis: the medians of incubation period between two groups are the same. The *p*-values for corresponding alternative hypotheses: the age<40 group has a smaller incubation median is 0.00474. It suggests that the age<40 group has a shorter incubation period than the age>=40 group. Similarly, the Siegal-Tukey test indicates the younger group’s incubation variance is less than the older group’s by *p*-value: 0.0083. It may suggest COVID-19 has a faster but relatively constant spread speed among people <40 years old than people >=40 years old.

### The incubation data of COVID-19 is linearly separable for Age>=40 and Age<40 groups

Figure 4 illustrates biplots of the dataset by removing two cases with missing items by using PCA (principal component analysis), sparse PCA (sparse principal component analysis, t-SNE (t-distributed stochastic neighbor embedding), and LLE (locally linear embedding) [16-19]. Data is partitioned as the age>=40 and age<40 groups in visualization. t-SNE shows that only one case in the age>=40 group falls in the cluster of the age < 40 group. But PCA, sparse PCA and LLE all show that the incubation of two groups is linearly separable, which means there exists an obvious linear boundary to separate them, in the subspaces generated by PCA, SPCA, t-SNE, or LLE. Such machine learning results indicate that the two groups are actually independent clusters spatially. However, we also find that the incubation data will no longer demonstrate the linear separability property when we partition it as age>=50 and age<50 groups or age>=55 and age<55 groups. It suggests that age 40 can be a key age cutoff for the incubation of COVID-19 along with previous statistical analysis.

**Fig 4.**
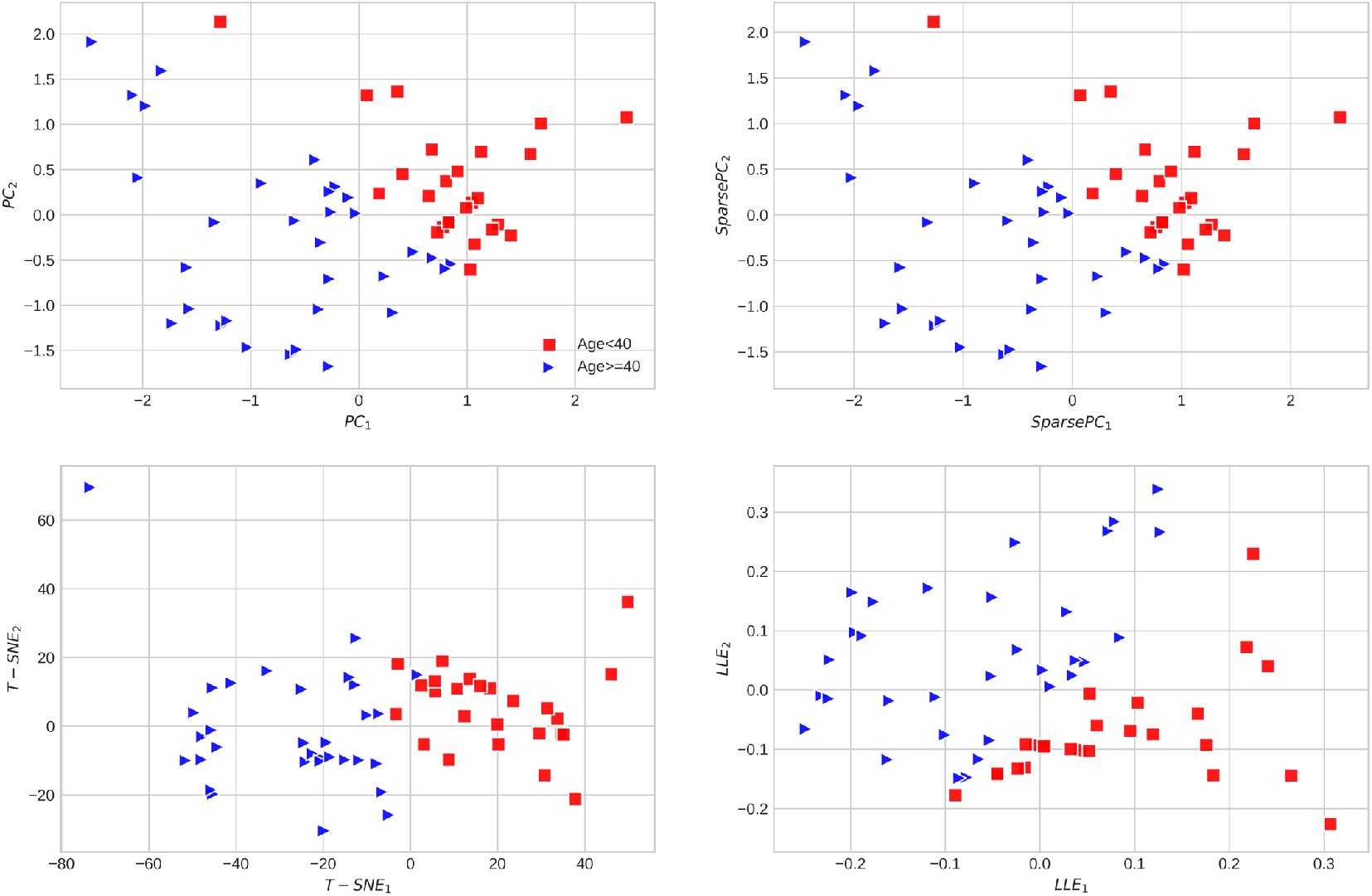
The PCA, sparse PCA, t-SNE, and LLE visualization of the incubation of COVID-19 partitioned as Age>=40 and Age<40 groups

### COVID-19 incubation statistics estimation

We further estimated population mean and standard deviation, median and 2.5^th^ and 97.5^th^ percentile by using Bootstrap under 10^6^ times resampling. Table 3 illustrates that their estimated results for COVID-19 along with the proposed Monte Carlo simulation estimation for the mean: Mean^MC^ and median: Median^MC^. The median and mean estimations from Monte Carlo simulation are consistent with those of bootstrap. As such, we believe the estimated median and mean of COVID-19 are 5.0 and 5.84 days. We still choose the confidence interval estimations from the bootstrap approach as the final 95% confidence intervals because we choose a smaller variance control parameter in the Monte Carlo simulations. Figure 5 illustrates the histograms of incubation medians, means, and the size of large incubation samples in simulation, all of which are normally distributed.

**Table 3.** Estimated COVID-19 incubation statistics

| Methods | Estimation<br>(days) | 95% CI |
| --- | --- | --- |
| Mean | 5.84 | (5.07, 6.55) |
| Mean<sup>MC</sup> | 5.84 | (5.42, 6.25) |
| Std | 2.98 | (2.31, 3.72) |
| Median | 5.01 | (4.00, 6.00) |
| Median<sup>MC</sup> | 4.99 | (4.59, 5.39) |
| 2.5 percentile | 2.69 | (2.45, 2.95) |
| 97.5 percentile | 12.89 | (11.00, 16.13) |

**Fig 5.**
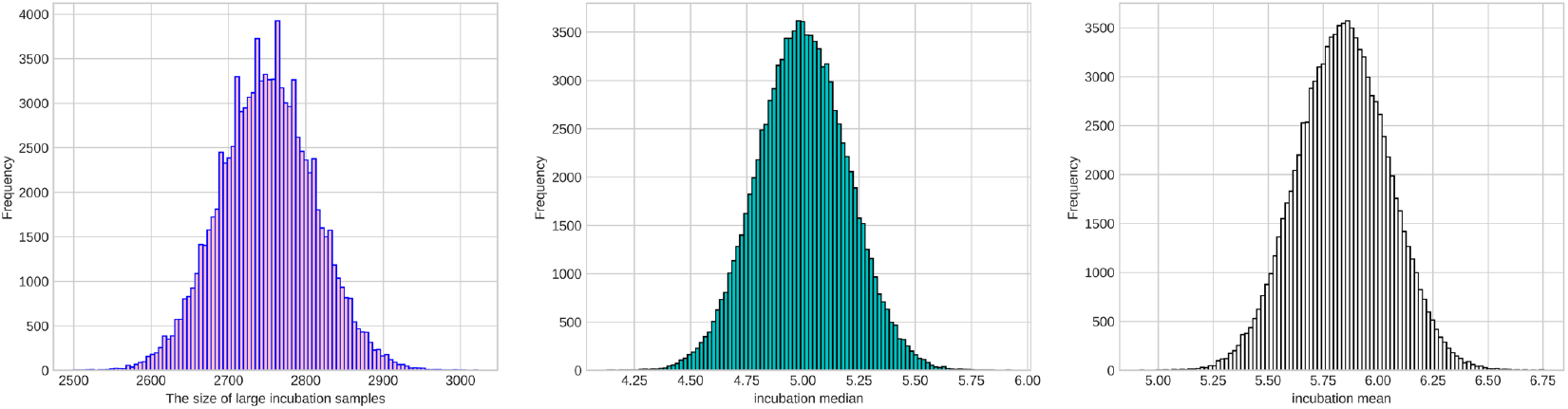
The histograms of the sizes of large samples, incubation median, and means in simulation.

## Discussion

We estimate the incubation period of COVID-19 in this study and analyze its properties by employing statistical and machine learning techniques. We estimate that the mean and median of COVID-19 as 5.84 and 5.01 days by employing nonparametric techniques. The proposed Monte Carlo simulation method echo the nonparametric estimation results. The incubation median of COVID-19: 5.01days (95% CI (4.0, 6.0) days is close to that of SARS (4.6 days) (95%CI: (3.8,5.8)). Compared to the incubation mean of MERS: 5.5 days (95% CI: (3.6-10.2) days), the COVID-19 mean incubation period: 5.84 days (95% CI: (5.07, 6.45)) are also slightly longer [21]. It suggests that COVID-19 could have a faster distribution speed than H7N9, but the same spread speed as SARS and MERS in terms of their incubation periods [22]. The existing spread of COVID-19 is faster than SARS partially because it has more complicate spread dynamics [2]. For example, those without clinical symptoms can still spread the virus even if they are ‘officially negative’ in the COVID-19 virus test [22].

We also investigate the incubation period of 12 family cases and 47 non-family cases in the dataset. The family cases simply refer to the patients who were caught by COVID-19 because their family members had been infected. The Mann-Whitney rank test shows that there are not significant differences between family patients and non-family patients in terms of the median of incubation period. The Siegal-Tukey tests on scale also verify that the incubation scales are at the same level for family patients and non-family patients.

Our studies indicate that incubation periods of the age>=40 years and age<40 years groups not only statistically significant but also linearly separable in machine learning. It may suggest different treatments should be considered for the two different groups. It will be more interesting to estimate different incubation time for them separately. That the estimated 97.5^th^ percentile of COVID-19 incubation is 12.89 days (95% CI: (11.00, 16.13)) may suggest a long isolation or quarantine time (e.g. 17 days) can be better than the widely accepted 14 days. Furthermore, different quarantine time should be applied to the age>=40 years and age<40 years groups for their different incubation periods. Generally speaking, a longer quarantine time can be considered for the old patients (>=40 years) than young patients (<40 years old). Our ongoing work is to collect more qualified data to extend our existing results and investigate incubation of COVID-19 for different groups besides comparing our incubation estimation with other studies [23].

## Data Availability

https://github.com/hank08819/COVID-19

## Acknowledgement

This research was funded by The Emergency Program for Prevention and Control of Novel Coronavirus of Xi’an Medical University (grant No. 2020ZX01).

